# TIME TO LUNG VOLUME STABILITY AFTER PRESSURE CHANGE DURING HIGH-FREQUENCY OSCILLATORY VENTILATION

**DOI:** 10.1101/2021.01.28.21250723

**Authors:** David G Tingay, Nicholas Kiraly, John F Mills, Peter A Dargaville

**Affiliations:** Neonatal Research, Murdoch Children’s Research Institute, Parkville, Australia; Department of Neonatology, Royal Children’s Hospital, Parkville, Australia; Department of Paediatrics, University of Melbourne, Melbourne, Australia; Department of Paediatrics, Royal Hobart Hospital and University of Tasmania, Hobart, Australia

**Author notes:** **Corresponding Author**: A/Prof David Tingay MBBS FRACP PhD, Neonatal Research, Murdoch Children’s Research Institute, 50 Flemington Rd., Parkville 3052 Victoria, Australia. **Author Contributions**: DGT, PAD, JFM developed the concept and designed the study. DGT enrolled and studied all infants. NK developed the mathematical models used in the study. NK, DGT and PAD were involved in data analysis and interpretation. All authors contributed to drafting the final manuscript with NK and DGT writing the first draft. **Category of Study**: Clinical (observational). Consent Statement: Prospective written informed parental consent was obtained for each infant prior to enrolment in the study. **Data Sharing**: Individual participant data collected during the study, after de-identification, and study protocols and statistical analysis code are available beginning 3 months and ending 23 years following article publication to researchers who provide a methodological sound proposal, with approval by an independent review committee (“learned intermediatry”) identified for purpose. Data is available for analysis to achieve aims in the approved proposal. Proposals should be directed to; to gain access, data requestors will need to sign a data access or material transfer agreement approved by the Murdoch Children’s Research Institute.

**Keywords:** High frequency oscillatory ventilation, infant, mechanical ventilation, lung mechanics

## Abstract

**Objectives:** Clinicians have little guidance on the time needed before assessing the effect of a mean airway pressure (P_AW_) change during high-frequency oscillatory ventilation (HFOV). We aimed to determine 1) time to stable lung volume after a P_AW_ change during HFOV and, 2) the relationship between time to volume stability and the volume state of the lung.

**Methods:** Continuous lung volume measurements (respiratory inductive plethysmography) after 1-2 cmH_2_O P_AW_ changes made every 10 minutes during an open lung strategy (n=13 infants) were analysed with a bi-exponential model. Time to stable lung volume (extrapolated to maximum 3600s) was calculated if the model R^2^ was >0.6.

**Results:** 196 P_AW_ changes were made, with no volume change in 33 (17%) occurrences. 125 volume signals met modelling criteria for inclusion; median (IQR) R^2^ 0.96 (0.91, 0.98). The time to stable lung volume was 1131 (718, 1959)s (P_AW_ increases) and 647 (439, 1309)s (P_AW_ decreases), with only 17 (14%) occurring within 10 minutes and time to stability being longer when the lung was atelectatic.

**Conclusions:** During HFOV, the time to stable lung volume after a P_AW_ change is variable, often requires more than 10 minutes and is dependent on the preceding volume state.

**Impact Statement:**

- In infants without preterm respiratory distress syndrome the time to achieve lung volume stability after a P_AW_ change during HFOV is usually greater than 10 minutes.
- The volume state of the lung at the time of P_AW_ change influences the time required to achieve a stable new lung volume; being shorter when the lung is well recruited and longer when the lung is already atelectatic.
- Clinicians should be aware that it may require least 10 minutes before assessing the clinical response to a change in P_AW_ during HFOV

## INTRODUCTION

The safe and effective delivery of high-frequency oscillatory ventilation (HFOV) depends on achieving an optimal lung volume.(1) During HFOV, the principal determinant of lung volume is the applied mean airway pressure (P_AW_).(2) Optimally applied, P_AW_ maximises oxygenation(3-5) and lung mechanics(6-8), whilst an inappropriate P_AW_ increases adverse events(9) and cardiovascular compromise(10) due to either atelectasis or overdistension. The most recent European guidelines on the management of preterm respiratory distress syndrome (RDS) recommend using an open lung strategy on initiation of HFOV.(11) Open lung strategies involve mapping the quasi-static pressure-volume relationship of the lung using a series of increasing, and then decreasing, P_AW_ steps applied over a fixed period of time, with the purpose of identifying the point of optimal oxygenation upon the deflation limb of the pressure volume relationship of the lungs. Usually, peripheral oxygen saturation (SpO_2_) and inspired oxygen fraction (FiO_2_) are used to guide the response.(3, 12, 13) Critical to the practical application of open lung strategies is an understanding of how rapidly lung volume stabilises after each P_AW_ change.

Due to the non-linear mechanical properties of the respiratory system, changes in lung volume follow an exponential plateau pattern after an adjustment in P_AW_.(14-16) The time required to achieve a new steady state lung volume is determined by the time constant of the respiratory system. An understanding of this relationship has important clinical implications for the application of open lung strategies during HFOV. Adequate time must be allowed for achievement of the desired new lung volume with changes in SpO_2_ reflecting the new volume.(7) In preterm infants with early RDS receiving HFOV as first intention invasive respiratory support, lung volume stabilises within 5 minutes of a 2 cmH_2_O P_AW_ change, and varies based on the volume state of the lung; well recruited lungs require less time than partially recruited lungs.(17) Most infants receiving HFOV do not have acute RDS, and many are not preterm.(18) Thus, the poor lung compliance and short time constants associated with acute RDS in preterm infants are unlikely to be translatable to the majority of infants being managed with HFOV as rescue therapy.

We aimed to determine: (1) time to stable lung volume after a P_AW_ change during HFOV and, (2) the relationship between time to stable lung volume and the volume state of the lung. These aims are addressed by applying different exponential models of the volumetric behaviour of the lung against time from a series of infants we have previously reported as part of other studies, in which the clinical and volume response to an open lung strategy was measured.(3, 8)

## METHODS

This study was performed at the Neonatal Unit of the Royal Children’s Hospital, Melbourne and was approved by the Royal Children’s Hospital Human Ethics Committee. Prospective, written, informed parental consent was obtained for each infant prior to enrolment in the study. The dataset used for this study was recorded from infants studied from 2004 to 2006. A detailed methodology has been published previously.(3, 8)

### Study Population

Infants receiving HFOV, using the Sensormedics 3100A oscillator (Sensormedics, Yorba Linda, CA), and muscle-relaxants were studied. Infants were not eligible if they had congenital heart disease, a known chromosomal anomaly, refractory hypotension or an inspired oxygen fraction (F_IO2_) of greater than 0.9.

### Measurements

Proximal P_AW_ was measured at the airway opening using a Florian respiratory monitor (Acutronic Medical Systems, Zug, Switzerland). Real-time relative changes in lung volume were measured with a low-pass-filtered, DC-coupled respiratory induction plethysmograph (RIP; Respitrace 200™, Non-invasive Monitoring Systems Inc., North Bay Village, FL) using the technique we have described previously to derive an uncalibrated volume signal in volts,(3, 19) following signal thermal stability.(20)

The pressure-volume relationship of the lung was mapped in all infants as part of another study.(3) To summarise this protocol: after achieving an F_IO2_ that maintained a stable SpO_2_ of 90-94%, a series of 2 cm H_2_O P_AW_ increases were made every 10 min (inflation limb) from the P_AW_ in clinical use (P_initial_) until no further improvement in SpO_2_ was noted over two consecutive P_AW_ increments (P_max_; functional total lung capacity). The deflation limb was then mapped by decreasing P_AW_ in 1-2 cm H_2_O steps every 10 min (deflation limb) until the P_AW_ was identified that resulted in SpO_2_ <85% for 5 min (P_final_; closing pressure of the lung).(3, 4, 21)

### Data collection and analysis

P_AW_ and V_RIP_ were recorded at 200 Hz, digitized and analysed using custom-built software (LabVIEW™, National Instruments, Austin, TX). From each RIP recording, the amplitude (V_TRIP_; Volts) and trough of each tidal oscillation was determined, with the trough voltage defining end-expiratory volume (V_LRIP_). P_AW_ and V_LRIP_ were normalized to the values at P_max_ (100%) and P_final_ (0%).(3) Initially, the time course of the V_LRIP_ signal was analysed to determine if any volume change occurred within the 10 min period (Figure 1). A detectable change was defined as a difference between initial and final V_LRIP_ voltage of at least 1/3 of the average oscillatory amplitude (V_TRIP_ value) at that P_AW_. This definition was chosen to account for the facts that RIP cannot be reliably calibrated to a known volume during HFOV, has a 3-6 % measurement error, and that V_TRIP_ would represent 1-3 mL/kg.(22)

**Figure 1.**
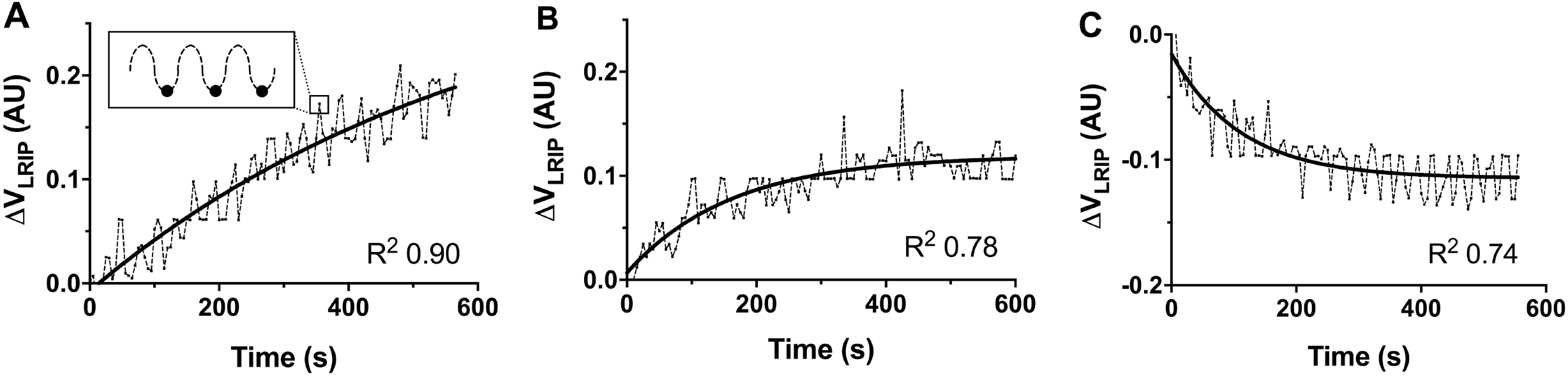
Representative examples of lung volume changes (ΔV_LRIP_, Arbitrary Units; AU) over 10 min following a single adjustment in P_AW_ of 2 cmH_2_O during the Inflation (**A** and **B**) and Deflation (**C**) limb. Dotted line: V_LRIP_, plotted using sequential trough values from the oscillatory waveform (see inset; black dots indicate trough of oscillatory V_LRIP_ time course); solid line: fitted bi-exponential model. In **Panel B** and **C** stable lung volume was achieve before 600s; 241s (R^2^ 0.78) and 168s (R^2^ 0.74), respectively. In panel A, stable lung volume was not achieved by 600s.

In the recordings in which V_LRIP_ did change, a second-order bi-exponential model was applied to the time course signal:(23)

### Inflation Limb

*y* = *y*_0_+ a(1 – *e*^-*t*/*τ*1^) + b(1 – *e*^-*t*/*τ*2^)

### Deflation Limb

*y* = *y*_*f*_+ a.*e*^-*t*/*τ*1^ + b.*e*^-*t*/*τ*2^

where *y* is V_LRI*P*_, *y*_0_ is initial V_LRIP_ for each time signal recording and *y*_*f*_ final V_LRIP_, *t* is time since P_AW_ change(s), *a* and *b* define the magnitude of volume such that final V_LRIP_= *y*_0_+ *a* + *b*, and τ_1_ and τ_2_ are time constants.

This model is superior to other non-linear models in a population of spontaneously breathing adults with and without lung disease.(23) Using an extra sum-of-squares F test comparison against a simpler single-order exponential equation,(15) this model is valid in over 90% of recordings. The time to achieve stable lung volume (defined as 95% of total ΔV_LRIP_ predicted by the model) was only calculated from those V_LRIP_ data in which the model had a goodness-of-fit of R^2^ ≥0.6. If stability had not occurred within the 10 min recording period, the time extrapolation was permitted to a maximum of 3600 s. Statistical analysis was performed with Prism 9.0 (GraphPad, San Diego, CA, USA) and a p value <0.05 was considered significant.

## RESULTS

Thirteen term or near-term infants, corrected for gestational age, were studied. All infants completed the protocol without complications. Their demographic and clinical characteristics are summarised in Table 1. Seven infants were receiving HFOV to treat meconium aspiration syndrome, four infants had pneumonia and the remaining two infants required HFOV following abdominal surgery. One of these infants was ex-preterm and had evolving chronic lung disease.

**Table 1.** Subject characteristics at study commencement (n=13)

| Age<br>(days) | Weight<br>(kg) | GA<br>(weeks) | Time on HFOV<br>(hr) | Initial HFOV settings |  |  |  | Gas exchange indices |  |  |
| --- | --- | --- | --- | --- | --- | --- | --- | --- | --- | --- |
|  |  |  |  | P<sub>AW</sub><br>(cm H<sub>2</sub>O) | ΔP<br>(cm H<sub>2</sub>O) | Fr<br>(Hz) | F<sub>IO2</sub> | P<sub>aCO2</sub><br>(mmHg) | AaDO<sub>2</sub><br>(mmHg) | OI |
| 2 | 3.4 | 40 | 24 | 14.3 | 33 | 8 | 0.5 | 48 | 197 | 9.6 |
| (1, 42) | (1.1, 3.7) | (23, 42) | (4, 54) | (9.8, 17.4) | (21, 42) | (6, 12) | (0.21, 0.9) | (37, 100) | (35, 526) | (5.5, 31.6) |

All data Median and Range.
GA; gestational age, HFOV: high frequency oscillatory ventilation, ΔP; Amplitude, Fr; Frequency, P<sub>aCO2</sub>; Partial arterial pressure of carbon dioxide, AaDO<sub>2</sub>; alveolar-arterial oxygen difference, OI; oxygenation index

A total of 196 P_AW_ changes were made (54 inflation limb, 142 deflation limb; Supplementary Figure 1). During the deflation series, the P_AW_ decrements were of magnitude 2 cmH_2_O (n=41) or 1 cmH_2_O (n=101). The time for P_AW_ to stabilise after a change was 9 (2, 27) s [median (range)]. 163 (83%) P_AW_ changes resulted in a volume change that met the predefined ΔV_LRIP_ criterion for inclusion in the analysis. During the deflation series, a ΔV_LRIP_ satisfying the inclusion criterion was more likely following a 1 cmH_2_O change (82% vs 66%), as 1 cmH_2_O changes were made around P_final_ where volume state was less stable.(7, 8) The exponential model could be fitted to the V_LRIP_ data with a R^2^ >0.60 for 125 P_AW_ alterations; median (IQR) R^2^ 0.96 (0.91, 0.98).

Figure 1 shows examples of inflation and deflation limb recordings. Only 3 (6%) inflation limb and 14 (18%) deflation limb recordings (n=125) achieved stable V_LRIP_ within the 10 min recording time (p=0.067, chi-square test). Stabilisation time was predicted by extrapolation to be within 3600 s (60 min) in a further 66 (53%) recordings. Using the model, the median (IQR) time to stable lung volume was 1311 (718, 1959) s in the inflation limb, and 647 (439, 1309) s in the deflation limb (p=0.023, Mann-Whitney test). The time constant of the first phase of the bi-phasic exponential model (τ_1_) was median 8 (3, 21) s.

Figure 2 shows the frequency distribution of all ΔV_LRIP_, with 56% (inflation) and 31% (deflation) of all P_AW_ changes requiring at least 30 min until stable V_LRIP_ (both p<0.0001; chi-square). Stable ΔV_LRIP_ was more likely to be obtained quickly during the first third of P_AW_ changes, and the last third more likely to require more than 30 min during the deflation limb (p<0.0001; chi-square). Figure 3 summarises the time to stable ΔV_LRIP_ within different regions of the pressure-volume relationship.

**Figure 2.**
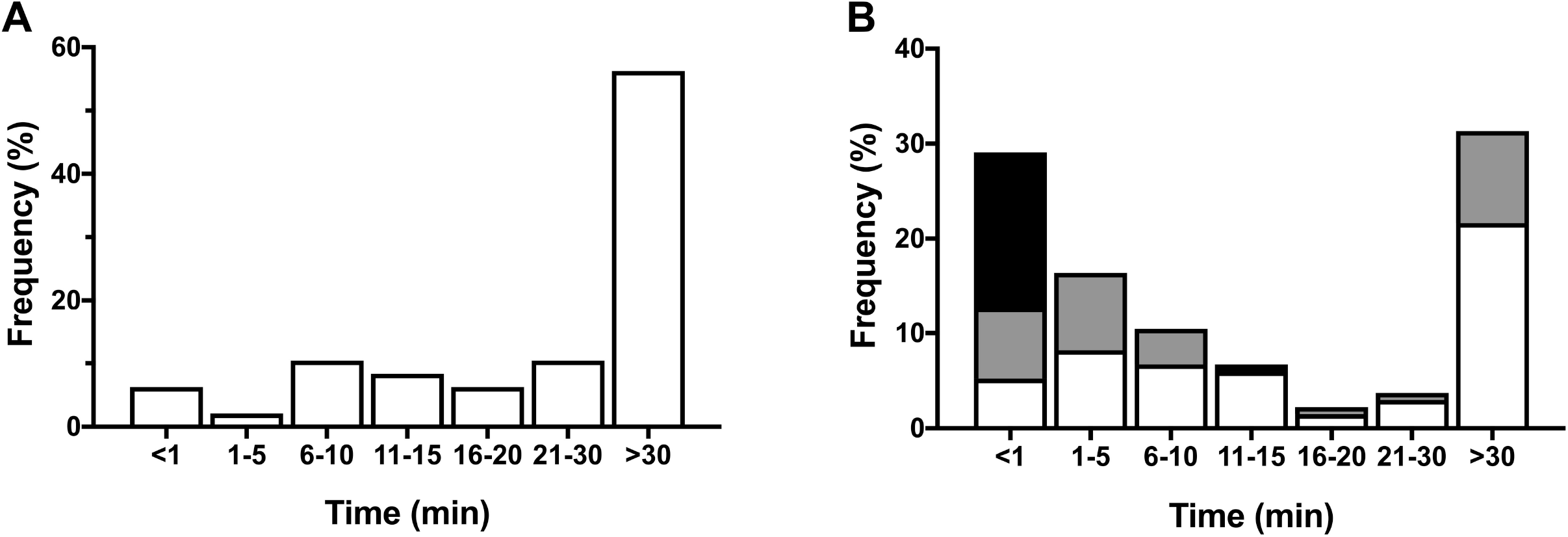
Frequency distribution of time to stable lung volume during the inflation (**A**; n=54) and deflation (**B**; n=142) limb P_AW_ changes. During the deflation limb, P_AW_ changes were analysed for the first third (black), middle third (grey) and last third (white) of sequential P_AW_ changes from P_max_ to P_final_. V_LRIP_ recordings that did not meet the criterion for volume change after the P_AW_ change are included in the <1 minute groups.

**Figure 3.**
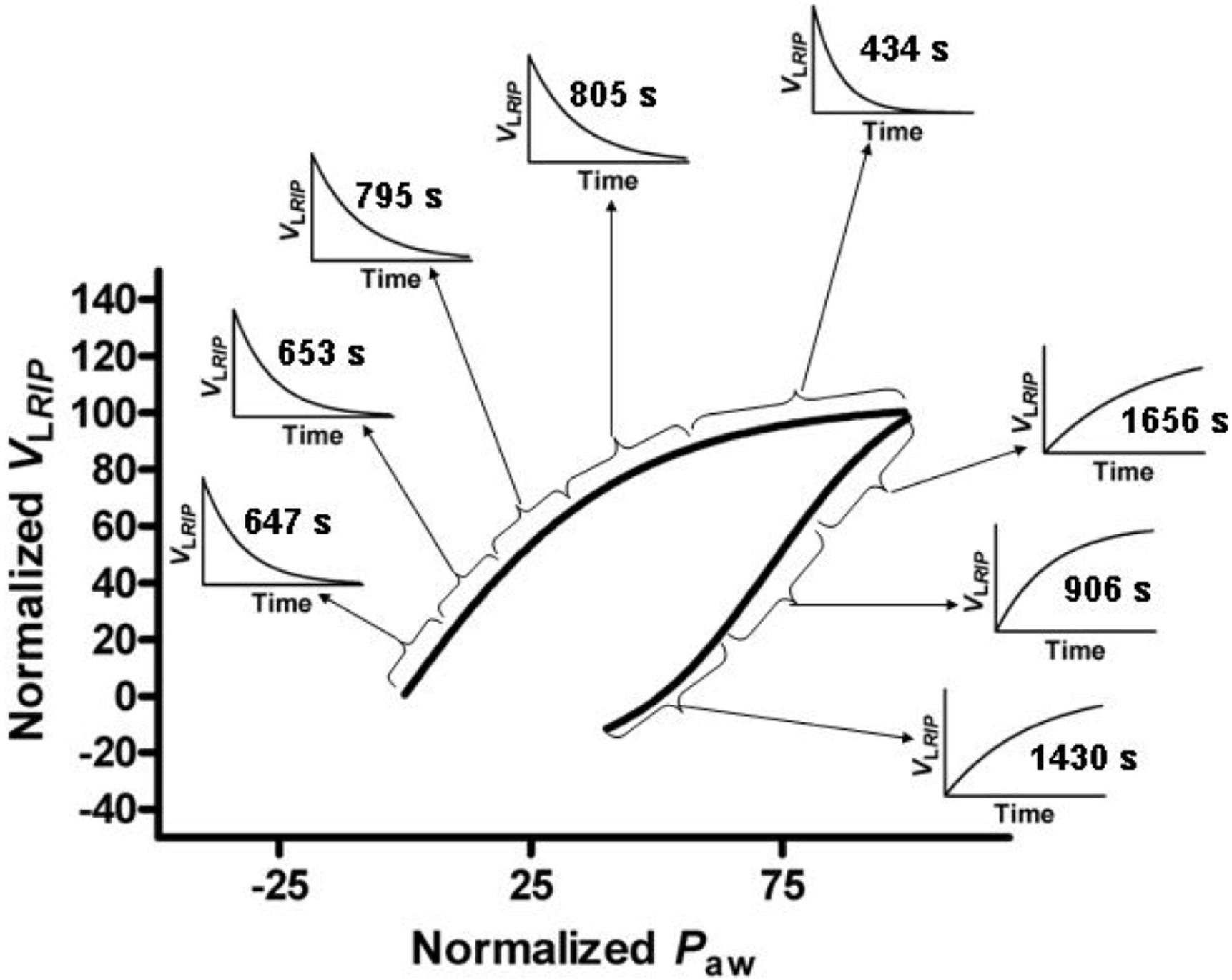
Schematic showing the median time to stable lung volume, and representative exponential model of lung volume change over time, for different sections of the normalised pressure-volume relationship of the lung measured during the study.(3) Regions of the pressure-volume relationship are separated by volume into equal thirds of the inflation limb and into equal fifths of the deflation limb.

## DISCUSSION

HFOV is used in the NICU for a diverse range of conditions,(18) and most often as a rescue therapy when conventional modes of mechanical ventilation are not effective. In our population of predominantly term infants receiving rescue HFOV, we found that lung volume had not fully stabilised within 10 min after a P_AW_ change in most cases. The time to stability was related to the volume state of the lung, reflecting that volume attainment is a continual and regional process, taking longer in poorly recruited lungs. These findings have implications for the application of open lung strategies during HFOV. Unlike the preterm infant with RDS, clinicians using HFOV in more mature infants will need to allow longer time before interpreting the clinical response to P_AW_ changes.

The importance of achieving an optimal lung volume during mechanical ventilation is well understood.(1, 2, 24, 25) During HFOV, P_AW_ is the principal determinant of lung volume, but clinicians have few guidelines on setting P_AW_. We found that the time to a stable lung volume after a 1-2 cmH_2_O P_AW_ change exhibited high inter- and intrasubject variability. The largest determinant of ΔV_LRIP_ was the volume state of the lung, with the time to stable V_LRIP_ being twice as long during the inflation limb compared to deflation limb. This was expected. During the deflation limb the lung was initially well recruited (‘open’), and HFOV occurred on the deflation limb of the pressure-volume relationship. On the deflation limb, alveoli are already recruited, and the lung is in a state of uniform volume. This creates alveolar stability and changes will be smaller and more rapid as long as the volume state remains above the closing pressure.(8, 26-28) During the inflation limb, recruitment is ongoing with some alveoli atelectatic, others recruiting or recruited, and some potentially overdistended. This heterogeneity of alveolar state, along with poorer lung compliance, increases time constants.(8, 26-28) Our data suggest that clinicians should consider the volume state of the lung when anticipating the clinical response to a P_AW_ change.

Only 14% of P_AW_ changes resulted in a stable lung volume within 10 minutes, with 42 changes (34%; 21 inflation and 37 deflation limb) predicted to require more than 60 minutes to stabilise. In contrast, a similar study in spontaneously breathing preterm infants receiving an open lung approach to HFOV for acute RDS reported stable lung volumes (measured with real-time continuous electrical impedance tomography; EIT) within approximately 5 min of a P_AW_ change.(17) Thome and co-workers reported a large time range of 2 to 25 min (median 9 min) for lung volume stabilisation in preterm infants following P_AW_ changes.(29) In this study lung volume was measured intermittently with the SF_6_ washout technique.(29) This required temporary conventional ventilation for 1 minute, which may have influenced the findings. Our study involved larger and more mature infants, all of whom were receiving rescue HFOV. None of the infants had primary RDS but rather pathologies analogous to acute respiratory distress syndrome.(30) Absolute lung volume, resistance and compliance is likely to be greater than preterm infants with RDS. With the recent recommendation to use an open lung approach on initiation of HFOV in preterm RDS,(11) clinicians must be aware that the recommended 2-3 minute P_AW_ step changes cannot be extrapolated to other neonatal respiratory conditions.(5, 12)

We applied a biphasic exponential model to describe volume change within the lung over time, the first time it has been applied to mechanically ventilated patients. This model has been previously used to describe forced expiration manoeuvres in adults(23) and passive expiration in newborn lambs.(31) In our population, the biphasic exponential model described the V_LRIP_ data well, and allowed extrapolation beyond the 10 min P_AW_ application period to predict ΔV_LRIP_ behaviour. This model allows for an initial rapid phase of volume change followed by a prolonged slow phase, generating a specific time constant for each. The model was consistent with the raw time-V_LRIP_ recordings and is biologically plausible. Open airways and alveoli have a direct connection to large airways and are therefore expected to change volume rapidly after a change in pressure, explaining the very rapid time constants observed in the first phase of the model. However, alveolar opening (recruitment) and closing (de-recruitment) occur more slowly, and are unpredictable,(32) especially in the presence of the noise associated with higher frequency pressure changes within the airways.(33)

Open lung approaches often report an initial increase in lung volume following reductions in initial P_AW_ from functional total lung capacity at P_max_.(3, 4, 34) We, and others, have postulated that this unexpected observation reflects the opening of small airways that were compressed, or release of impeded venous return, at higher P_AW_.(3, 4, 34) The bimodal model offers a third possibility that the slow phase of alveolar recruitment is still ongoing during the initial phase of the deflation limb. Whilst the lung is mechanically stable (above closing pressure) on the deflation limb, the initial V_LRIP_ reductions reflect open lung units rapidly achieving a stable volume, with the slower recruitment of incompletely opened lung units during the previous P_AW_ steps, with longer time constants, occurring in the background.

This study is limited to secondary analysis of an existing data set of 13 diverse infants. Although similar in size to previous reports in preterm infants,(4, 17) the infants in our study were receiving muscle relaxants. It is likely that spontaneous breathing will shorten the time to stable volume but also create more uncertainty in the response. Measuring lung volume change in infants during HFOV is difficult. Methods validated in other populations, such as inert gas washout,(29) chest radiograph(35) and computerised tomography,(36) are impractical, intermittent or involve unacceptable radiation. DC-coupled RIP is a well-validated, non-invasive, radiation-free method of continuously monitoring thoracic volume change during HFOV in animals(37) and infants,(3, 8, 19, 38) but is not without some limitations.(3) In particular, RIP cannot differentiate between gas and fluid changes within the chest, unlike newer technologies such as EIT.(39) EIT is also able to define regional volume states.(40) Unfortunately practical EIT systems were not available at the time this study was performed.

## Conclusions

In term and older infants receiving rescue HFOV, the time to a stable lung volume after a P_AW_ change is variable, and shorter when the lung is already recruited compared to when it is de-recruited. Unlike preterm infants, greater than 10 minutes is often required for lung volume stability. Clinicians should be aware of the importance of the preceding volume state in the lung when assessing the response to a P_AW_ change, and the need to allow longer for bedside monitoring to achieve a clinical response.

## Data Availability

Individual participant data collected during the study, after de-identification, and study protocols and statistical analysis code are available beginning 3 months and ending 23 years following article publication to researchers who provide a methodological sound proposal, with approval by an independent review committee (learned intermediatry) identified for purpose. Data is available for analysis to achieve aims in the approved proposal. Proposals should be directed to; to gain access, data requestors will need to sign a data access or material transfer agreement approved by the Murdoch Childrens Research Institute.

## Acknowledgements

The authors wish to acknowledge Prof Colin J Morley MD, who supervised DGT during the PhD work that generated the dataset used for this analysis, for his mentoring and advice.

