## Supplementary Figure 1 for "TIME TO LUNG VOLUME STABILITY AFTER PRESSURE CHANGE DURING HIGH-FREQUENCY OSCILLATORY VENTILATION"

**Supplementary Material**

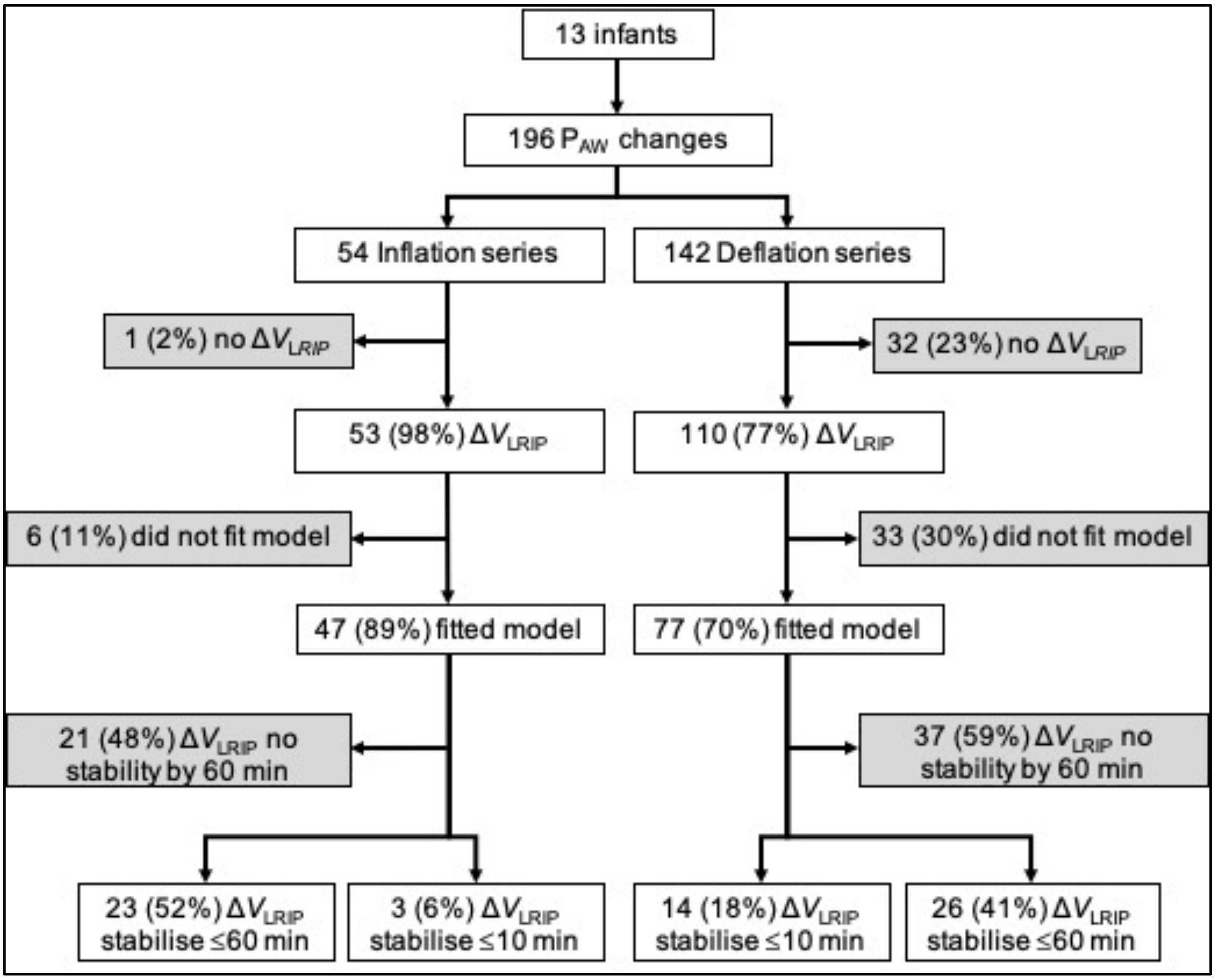


**Supplementary Figure 1.** Study analysis flow chart
